# A map of the historical spread of *Cyclospora cayetanensis* with a focus on the United States of America (USA)

**DOI:** 10.64898/2026.08.01.26359445

**Authors:** Daniel Janies, Sayal Guirales-Medrano, Ana Clecia Dos Santos Silva, Colby T. Ford

**Affiliations:** Center for Computational Intelligence to Predict Health and Environmental Risks (cipher.charlotte.edu), The University of North Carolina at Charlotte, USA

## Abstract

*Cyclospora cayetanensis* causes significant seasonal foodborne illness, characterized by recent large-scale nationwide outbreaks. We are currently experiencing the largest recorded outbreak in USA history during the summer of 2026. The summer 2026 outbreak emphasizes the urgent need for enhanced surveillance of infectious diseases. In this study, we use phylogenetic network analysis via the StrainHub framework to reconstruct the historical spread of *C. cayetanensis* in the USA using only mitochondrial sequence data and place of isolation metadata collected from 1997 to 2022. By quantifying parasite importation to localities through the metric indegree centrality, we identify Texas as a primary sink for *C. cayetanensis* introduction. Our finding aligns with historical epidemiologic investigations such as the 2013 multistate outbreak. While our model demonstrates the efficacy of reconstructing transmission pathways from molecular data alone, it also reveals critical surveillance gaps, particularly the masking of geographic origins in public datasets and the current absence of molecular data for the 2026 event. Moving forward, integrating real-time molecular surveillance with robust network analysis is essential to shift from reactive to proactive intervention strategies. We conclude that improved data sharing and reporting across food, clinical, and farm sectors are vital for disrupting transmission cycles of parasites. This work will protect public health and ensure a fresh clean supply of key foods for the nutrition of Americans.

## Introduction

*Cyclospora cayetanensis*, an obligate intracellular parasite of the phylum Apicomplexa, is the causative organism of cyclosporiasis. Cyclosporiasis is an intestinal illness with protracted symptoms including diarrhea. *C. cayetanensis* is a foodborne and waterborne parasite and occurs in human populations worldwide (Dubey et al., 2022). Humans serve as the only known host for *C. cayetanensis*.

*C. cayetanensis* requires an obligatory environmental maturation phase. Newly shed oocysts are unsporulated and non-infectious. The oocysts require 7 to 14 days under favorable environmental conditions to mature into an infectious state termed sporulated oocysts [Centers for Disease Control and Prevention (CDC), 2025]. Once sporulation is complete, the oocyst contains infectious sporozoites. If a person ingests food or water contaminated with sporulated oocysts, the parasite can excyst, releasing its sporozoites in the gastrointestinal tract, causing infection (Almeria et al., 2019).

This biological requirement largely eliminates direct human-to-human propagation, shifting the primary vehicles of transmission to environmental sources. These sources are predominantly agricultural water and fresh produce that is harvested close to or in the ground, such as leafy greens, herbs, berries, and green onions (CDC, 2025).

The public health burden of *C. cayetanensis* is particularly pronounced during large-scale seasonal outbreaks. Contaminated irrigation, washing, or processing water serves as a key pathway for broad agricultural contamination (Naganathan et al., 2022), allowing contaminated produce to enter wide distribution networks prior to clinical detection.

Once contaminated food is ingested by a human, the incubation period ranges from 7 to 14 days or more. The infection leads to symptoms characterized by prolonged watery diarrhea, abdominal cramping, nausea, fatigue, and substantial weight loss (CDC, 2025). This long incubation period makes epidemiological traceback to food sources very difficult, as a patient will have had many meals in a period of weeks.

The first-line therapeutic regimen, trimethoprim-sulfamethoxazole (TMP-SMX), is clinically effective. Diagnosis remains challenging as sporadic oocyst shedding often necessitates repeated stool sampling that is analyzed via specialized microscopy or polymerase chain reaction (PCR) assays (CDC, 2025).

The systemic threat posed by this organism is underscored by recent epidemiological surveillance. According to the CDC Health Alert Advisory released on July 14, 2026 (CDC, 2026a), domestic surveillance identified the start of the seasonal increase with symptom onset dates beginning as early as May 1, 2026. As of the end of July 2026, a major nationwide outbreak in the United States has accounted for 6,707 confirmed cases, 423 hospitalizations, and an estimated 11,500 suspected cases across more than 45 states (CDC, 2026b). This represents the largest recorded outbreak of *C. cayetanensis* in USA history.

However, the summer 2026 outbreak’s rapid growth, high case volume, and broad geographic extent suggest that multiple contaminated sources may be involved. Molecular surveillance data for the summer 2026 outbreak remain unavailable to the public at the time of writing.

Outbreaks of *C. cayetanensis* have occurred periodically in the USA. For example, in 2013, an outbreak centered in Texas and spread to 25 states. The USA Centers for Disease Control and Prevention investigated hundreds of cases across multiple states to link the parasite to fresh produce including salad mixes and cilantro (Abanyie et al., 2015). The farm or country origins of the 2013 outbreak centered in Texas were not disclosed to the public.

Cyclosporiasis is a notifiable disease in the USA. Robust surveillance, early clinical reporting, treatment of patients, and improved agricultural intervention protocols are key to disrupting transmission cycles of *C. cayetanensis*. To better understand how *C. cayetanensis* disseminates in the USA, we examine the historical outbreaks of *C. cayetanensis* with a focus on the USA.

## Methods

Among several genetic loci pioneered for the phylogenetics of *C. cayetanensis*, researchers have advocated for the use of mitochondrial (mt) DNA (Barratt et al., 2022; Nascimento et al., 2019). These data are publicly available from the US National Institutes of Health’s GenBank (Sayers et al., 2020).

We focused on small rDNA and repeat regions, as many *C. cayetanensis* isolates have been sequenced for these loci and genetic variation is apparent. Isolation dates for *C. cayetanensis* for these loci span from 1997 to 2022 (supplemental files: data_on_isolation.txt; genbank_data_for_isolates.txt).

Many of the *C. cayetanensis* isolates in our dataset are associated with the aforementioned 2013 multistate outbreaks that mirror the conditions reported today. We also include isolates from other regions and outbreaks that are related by sequence to provide evolutionary and epidemiological context.

We developed mt genome backbone alignment with MAFFT (default and commands –adjustdirection) and used MAFFT (commands --addfragments --keeplength; Katoh et al., 2022) to align the small subunit sequences to the backbone. We curated a final alignment of 185 taxa and 6,319 aligned positions (supplemental file: genome-w-small-subunits-V6-trim.phy).

We masked leading and trailing gaps and performed a tree search with RAxML under the substitution model GTRGAMMA with the outgroup *C. cayetanensis* isolate CHN_HEN01 mitochondrion from Henan, China (version 7.4.2; supplemental file: raxml-7.4.2-pthreads-tcp.sh; Stamatakis and Ott, 2008).

We prepared a metadata file containing isolation locations and analyzed these data in concert with the RAxML tree using StrainHub (supplemental files: place_output_clean_mergefix.csv; RAxML_bestTree.genome-w-small-subunits-V6-trim.out; de Bernardi Schneider et al., 2020). We quantified the frequency of parasite importation into specific locations using indegree centrality (Freeman, 1979). Indegree centrality is a metric implemented in the StrainHub framework to determine the relative importance of nodes as destinations for metadata state transitions (in this case, the place of isolation). High indegree centrality indicates a location that experiences a high frequency of importation of parasites from other sources. Low indegree centrality indicates a location that rarely receives parasites from other nodes in the network.

## Results

In the case of these *C. cayetanensis* data and their places of isolation, we see the following patterns (supplemental file: StrainHub_indegree.csv; Figure 1).

**Figure 1.**
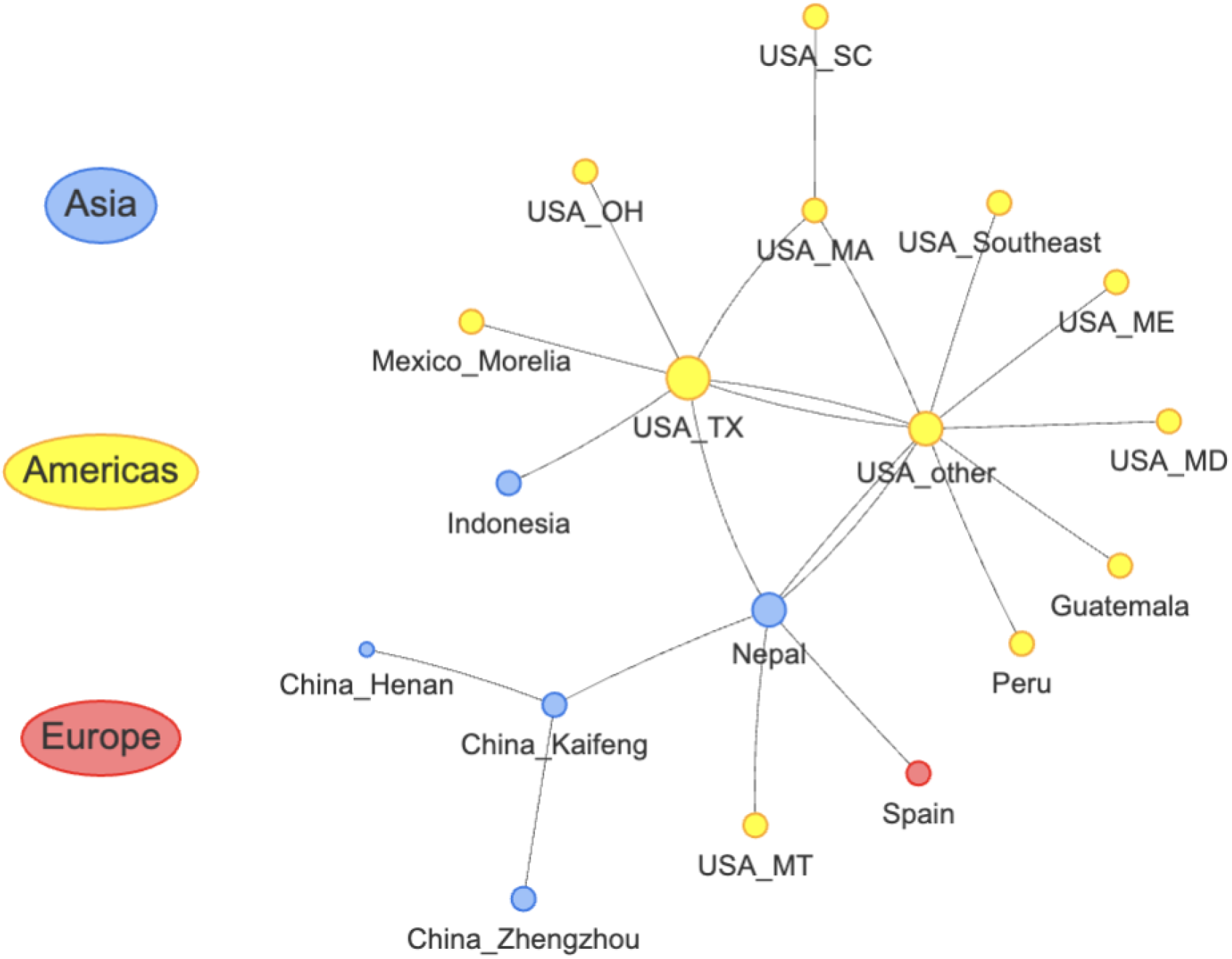
A StrainHub representing indegree centrality relationships between several isolates of *C. cayetanensis* and their places of isolation. Size of circles represents the magnitude of the indegree centrality. Colors represent large geographic designations. Abbreviations are as follows: TX = Texas, MA = Massachusetts, MD = Maryland, ME = Maine, MT = Montana, OH = Ohio, South Carolina = SC.

In Figure 1, we observe variable patterns of pathogen introduction across geographic regions as reflected by the indegree centrality metric (StrainHub_indegree.csv). This metric quantifies the number of directed transmission pathways converging on a specific geographic location, identifying it as a ‘sink’ or destination for parasite importation. USA Texas emerged as the primary sink in our model, recording the highest indegree centrality score (indegree = 3). Moderate levels of parasite importation (indegree = 2) were observed in the “USA other” (a designation used when reporting agencies mask specific geographic metadata) and Nepal. Other USA locations, including Massachusetts, Maryland, Maine, Montana, Ohio, South Carolina and the rest of the Southeast, had low degrees of importation (indegree = 1). International localities in these data including: China’s Kaifeng and Zhengzhou provinces, Guatemala, Indonesia, Morelia Mexico, Peru, and Spain also had low degrees of importation (indegree = 1).

As the locality of the outgroup taxon, China’s Henan province had an indegree centrality of 0. This is a condition of the underlying phylogenetic analysis needed to establish an evolutionary baseline (Maddison W. et al., 1984). We chose the isolate from Henan because it had been sequenced for a full mt genome and was indicated by Barrat et al., (2022) to be a distinct lineage (C) from the American lineages (A). With more data from more localities from related isolates, the traffic of parasites can be elucidated with these methods (Ezeoke et al., 2018).

## Discussion

The analysis of these data for *Cyclospora cayetanensis* transmission networks via indegree centrality highlights Texas as a primary sink for parasite introduction. The 2013 outbreak centered in Texas is etiologically similar to early reports of the 2026 summer outbreak involving food sources such as salad mixes (FDA, 2026). However, the summer 2026 outbreak may involve multiple food sources and foci.

The identification of Texas as a primary sink for *Cyclospora* importation in 2013 is supported by indegree centrality scores derived from phylogenetic network analysis. We used no literature hints up front to guide the data analyses.

Our analytical finding is consistent with the FDA’s July 2026 traceback investigation, which identified iceberg lettuce imported from Mexico as the source of the multistate outbreak (FDA, 2026). This result confirms that we can independently reconstruct epidemiologically supported investigations from only molecular sequences and place of isolation metadata.

The moderate levels of importation identified in regions such as Nepal, alongside specific domestic hubs, highlights the complex, multi-nodal nature of *C. cayetanensis* dissemination. While our phylogenetic framework provides a robust method for quantifying transmission pathways, it also exposes gaps in surveillance data. For example, the frequent use of just “USA” in public datasets masks the specific geographic origins of outbreaks, limiting the granularity of our analysis.

Furthermore, the absence of molecular surveillance data in the public domain for the 2026 outbreak limits our current ability to integrate the recent events into our model. We trust these data are forthcoming such that we can make a contribution to the fight against *Cyclospora cayetanensis*.

Moving forward, integrating real-time molecular surveillance with network analysis is essential to shift from reactive to proactive intervention. Moreover, with data from food items, the clinics, and the farms, we can create precise maps that will allow targeted interventions. These precise maps will afford better food safety without undue limits on commerce and consumption of clean food. Improved data transparency and standardized reporting will not only enhance our understanding of historical spread but will be vital for disrupting the transmission cycles of *C. cayetanensis* in the future and keeping the USA food supply safe.

## Data Availability

All data, code, and supplementary analyses are available on GitHub at:

https://github.com/janieslab/cyclospora_historical_spread_july2026

## Data and Code Availability

All data, code, and supplementary analyses are available on GitHub at: https://github.com/janieslab/cyclospora_historical_spread_july2026.

